# Substitutions in Spike and Nucleocapsid proteins of SARS-CoV-2 circulating in South America

**DOI:** 10.1101/2020.06.02.20120782

**Authors:** Carlos Franco-Muñoz, Diego A. Álvarez-Díaz, Katherine Laiton-Donato, Magdalena Wiesner, Patricia Escandón, José A. Usme-Ciro, Nicolás D. Franco-Sierra, Astrid C. Flórez-Sánchez, Sergio Gómez-Rangel, Luz D. Rodríguez-Calderon, Juliana Barbosa-Ramirez, Erika Ospitia-Baez, Diana M. Walteros, Martha L. Ospina-Martinez, Marcela Mercado-Reyes

## Abstract

SARS-CoV-2 is a new member of the genus *Betacoronavirus*, responsible for the COVID-19 pandemic. The virus crossed the species barrier and established in the human population taking advantage of the spike protein high affinity for the ACE receptor to infect the lower respiratory tract. The Nucleocapsid (N) and Spike (S) are highly immunogenic structural proteins and most commercial COVID-19 diagnostic assays target these proteins. In an unpredictable epidemic, it is essential to know about their genetic variability. The objective of this study was to describe the substitution frequency of the S and N proteins of SARS-CoV-2 in South America. A total of 504 amino acid and nucleotide sequences of the S and N proteins of SARS-CoV-2 from seven South American countries (Argentina, Brazil, Chile, Ecuador, Peru, Uruguay, and Colombia), reported as of June 3, and corresponding to samples collected between March and April 2020, were compared through substitution matrices using the Muscle algorithm in MEGA X.

Forty-three sequences from 13 Colombian departments were obtained in this study using the Oxford Nanopore and Illumina MiSeq technologies, following the amplicon-based ARTIC network protocol. The substitutions D614G in S and R203K/G204R in N were the most frequent in South America, observed in 83% and 34% of the sequences respectively. Strikingly, genomes with the conserved position D614 were almost completely replaced by genomes with the G614 substitution between March to April, 2020. A similar replacement pattern was observed with R203K/G204R although more marked in Chile, Argentina and Brazil, suggesting similar introduction history and/or control strategies of SARS-CoV-2 in these countries. It is necessary to continue with the genomic surveillance of S and N proteins during the SARS-CoV-2 pandemic as this information can be useful for developing vaccines, therapeutics and diagnostic tests.

**Highlights:**

- The spike and nucleocapsid proteins of SARS-CoV-2 circulating in Colombia and South-American countries have similar patterns of non-synonymous substitutions
- Substitutions D614G in Spike and R203K-G204R in Nucleocapsid are the most frequent in Colombia and South-American countries
- The identification of genetic variability of SARS-CoV-2, is useful for vaccines, diagnostic test, and therapeutic designs.

## 1. Introduction

The recently emerged SARS-CoV-2 responsible for the coronavirus disease 2019 (COVID-19) pandemic, has increased significantly in the number of cases and deaths, so that daily, about 70,000 new cases are reported globally (WHO, 2020a, 2020c). The first case of COVID-19 in South America was reported in Brazil on February 26, in a 61 years old man traveling from Italy (gob.br, 2020). In Colombia, the first case of COVID-19 was announced on March 6, in a traveler from Italy, after which the number of patients has exceeded 43,000 and over 1500 deaths (INS, 2020).

The SARS-CoV-2 genome consist of a single, positive-stranded RNA (ssRNA[+]), with 29,903 nucleotides long. This virus has shown an extraordinary infectiousness and transmission capacity in the human population (He et al., 2020), even exploring other vertebrate species (Shi et al., 2020). The SARS-CoV-2 genome has nine open reading frames (ORFs); the first one, subdivided in ORF1a and ORF1b by ribosomal frameshifting, encodes the polyproteins pp1a and pp1ab which are processed into non-structural proteins involved in subgenomic/genome length RNA synthesis and virus replication. Structural proteins, Spike (S), Envelope (E), Membrane (M), and Nucleocapsid (N) are encoded in subgenomic mRNA transcripts within ORFs 2, 4, 5, and 9, respectively (SIB, 2020; Yount et al., 2005)

Spike protein, a type I membrane glycoprotein, is the most exposed viral protein recognized by the cellular receptor angiotensin-2-converting enzyme (ACE2) during the infection of the lower respiratory tract and considered the main inducer of neutralizing antibodies. The N protein is associated with the RNA genome to form the ribonucleocapsid and is abundantly expressed during infection. Both N and S proteins are highly immunogenic and most commercial COVID-19 diagnostic tests (molecular and immunologic) target these proteins (Álvarez-Díaz et al., 2020; Lee et al., 2020).

Furthermore, non-synonymous mutations in the S and N proteins have been reported, their implications in the potential emergence of antigenically distinct and/or more virulent strains remain to be studied, although it was reported that mutations in the receptor-binding domain (RBD) at the S protein of SARS-CoV-related viruses disrupt the antigenic structure and binding activity of RBD to ACE2 (Du et al., 2009) Similarly, how non-synonymous mutations could impact the antibody response and the specificity and sensitivity of serological tests for COVID-19 diagnosis is unknown. Thus, identifying variable sites at these proteins can provide a valuable resource for choosing the target antigens for the development of SARS-CoV-2 vaccines, therapeutics, and diagnostic tests (Du et al., 2009; Jacofsky et al., 2020). The objective of this study is to describe the frequency of substitutions in S and N proteins of SARS-CoV-2 in South America.

## 2. Materials and methods

### 2.1 Ethics

This work was developed according to the national law 9/1979, decrees 786/1990 and 2323/2006, which establishes that the Instituto Nacional de Salud (INS) from Colombia is the reference lab and health authority of the national network of laboratories and in cases of public health emergency or those in which scientific research for public health purposes as required. The INS is authorized to use the biological material for research purposes, without informed consent, which includes the anonymous disclosure of results. This study was performed following the ethical standards of the Declaration of Helsinki 1964 and its later amendments. The information used for this study comes from secondary sources of data that were previously anonymized and do protect patient data.

### 2.2 Patients and samples

Nasopharyngeal swab samples from patients with suspected SARS-CoV-2 infection were processed for RNA extraction using the automated MagNA Pure LC nucleic acid extraction system (Roche Diagnostics GmbH, Mannheim, Germany) and viral RNA detection was performed by real-time RT-PCR using the SuperScript III Platinum One-Step Quantitative RT-qPCR kit (Thermo Fisher Scientific, Waltham, MA, USA), following the Charité-Berlin protocol (Corman et al., 2020) for the amplification of the SARS-CoV-2 E (betacoronavirus screening assay) and RdRp (SARS-CoV-2 confirmatory assay) genes.

### 2.3 Complete genome sequencing of SARS-CoV-2 through NGS

NGS of SARS-CoV-2 from 43 patients was performed using the amplicon-based Illumina and Nanopore sequencing approaches, ARTIC network protocol (Quick, 2020a). Following cDNA synthesis with SuperScript IV reverse transcriptase (Thermo Fisher Scientific, Waltham, MA, USA) and random hexamers (Thermo Fisher Scientific, Waltham, MA, USA), a set of 400-bp tiling amplicons across the whole genome of SARS-CoV-2 were generated using the primer schemes nCoV-2019/V3 (Quick, 2020a).

SARS-CoV-2 specific oligonucleotides were used for the generation of amplicons by means of a high-fidelity DNA polymerase (Q5® High-Fidelity DNA Polymerase - (New England Biolabs Inc., UK, EB), in order to avoid the introduction of artificial substitutions.

### 2.4 Sequence Analysis

Reads were mapped to the Wuhan-Hu-1 reference genome (NC_045512.2) using BWA (Li et al., 2020) and BBmap (brian-jgi, 2020); then, assembled sequences were submitted to GISAID. Substitution matrices of nucleotides and amino acids of S and N proteins were generated from a multiple sequence alignment with the reference genome against the 43 assembled Colombian SARS-CoV-2 genomes (Table 1) using the Muscle algorithm (Edgar, 2004) in MEGA X (Kumar et al., 2016). Subsequently,461 SARS-CoV-2 sequences from South American countries, including Argentina, Brazil, Ecuador, Peru, Uruguay and other sequences from Colombia available on the GISAID, NCBI, and GSA databases were analyzed (Supplementary Table S1, and Supplementary Table S2).

**Table 1.**
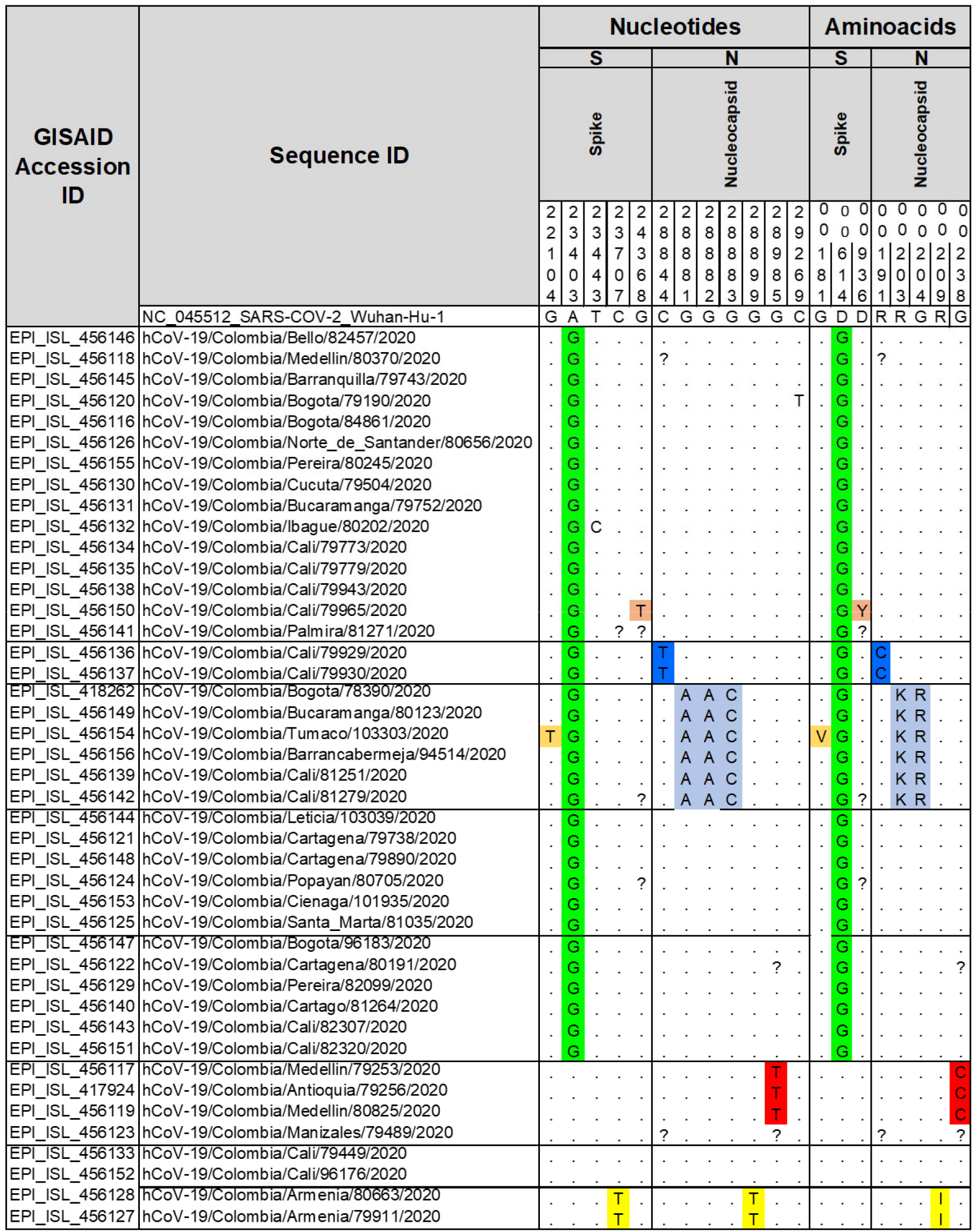
Nucleotide substitutions and amino acid changes in the Spike and Nucleocapsid proteins of 43 SARS-CoV-2 genomes from Colombia generated in this study. Location of the substitutions was estimated based on the reference genome NC_045512.2. Left panel: nucleotides; right panel: amino acids. ^?^: Uncovered nucleotide and amino acids during

## 3. Results

### 3.1 Non-synonymous substitutions in the Spike and Nucleocapsid proteins in Colombia

Several non-synonymous substitutions were observed in the S and N proteins of the Colombian SARS-CoV-2 sequences generated in this study. Three amino acid substitutions were observed in the S protein, D614G was present in 81% (35/43) of the sequences. Furthermore, substitutions G181V and D936Y were found in low frequencies of 2.3% (1/43) and 2.3% (1/43) respectively (Table 1). In the N protein, five amino acid substitutions were found; the most frequent being R203K and G204R in 13.95% (6/43) of the sequences. Amino acid substitutions, R191C, R209I and G238C were found in 4.65% (2/43), 4.65% (2/43) and 6.97% (3/43) of the Colombian sequences, respectively (Table 1). Some nucleotide substitutions were synonymous.

### 3.2 Non-synonymous substitutions in the Spike and Nucleocapsid proteins in South America

Genomic resource databases, NCBI, GISAID and GSA were consulted to determine the substitutions in S and N proteins of SARS-CoV-2 from South America. A total of 504 genomes reported as of June 3^Th^ 2020, were analyzed, 126 from Colombia (including the 43 genomes reported in this study), 29 from Argentina, 145 from Brazil, 153 from Chile, 4 from Ecuador, 2 from Peru and 45 from Uruguay. Fifty sequences of S and 27 of N were excluded from the analysis because the presence of undetermined bases that did not allow the proper identification of the S and N ORFs in the amino acid substitution matrices. Twenty-eight and twenty-two non-synonymous substitutions were identified in the sequence of S and N proteins respectively, in genomes of South America (Table S1 and S2). The most frequent in S were D614G (83%) V1176F (2.2%) and P1263L (1.5%), while the most frequent in N were R203K (34.5%), G204R (34.3%), I292T (15.8%) and S197L (3.3%). The remaining substitutions in both, S and N occurred in less than 1% of the sequences. These included G181V and D936Y in S, and R191C and G238C in N, as observed in the Colombian genomes (Fig. 1).

**Figure 1.**
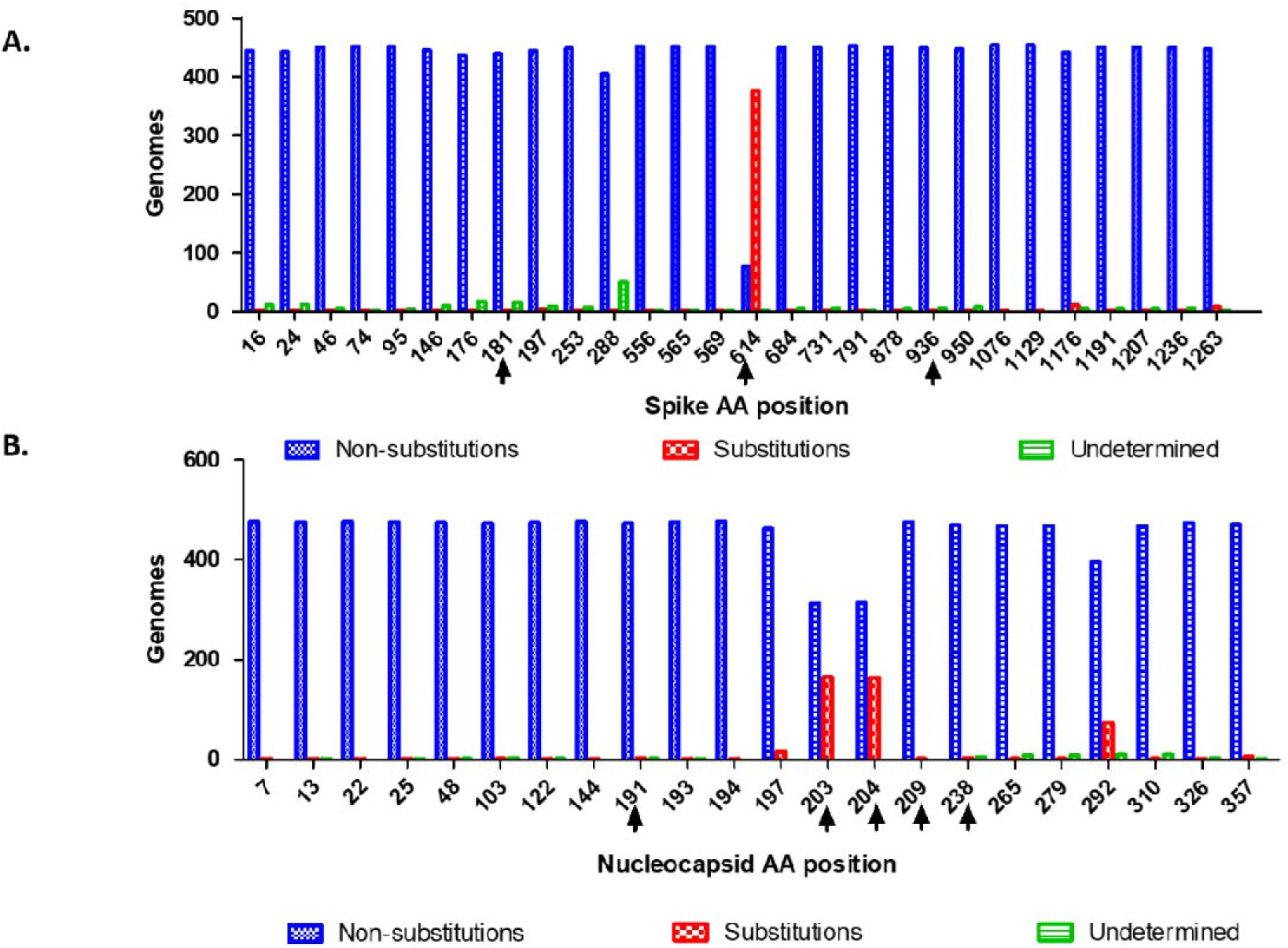
Non-synonymous substitutions in the Spike and Nucleocapsid proteins of SARS-CoV-2 in South America. **A**. Non-synonymous substitution sites in the S protein. Blue bars represent the genomes with conserved positions, red bars represent the genomes with substitution at the indicated site (at least one genome per site). **B**. Non-synonymous substitutions in the N. Blue bars represent the genome with conserved positions, red bars represent the genomes with substitution at the indicated site (at least one genome). Green bars represent the number of genomes with undetermined amino acids at the indicated site. Black arrows show substitutions found in the 43 Colombian genomes generated in this study. The location of the substitutions was estimated based on the reference genome NC_045512.2

### 3.3 Spatiotemporal distribution of substitutions in Spike and Nucleocapsid

The analysis of substitution frequencies by country shows that D614G substitution in the S protein was frequent in Argentina, Brazil, Chile, Colombia and Peru, with 80-100% of the reported sequences (Fig. 2A). In Ecuador and Uruguay D614 position was predominant by March, however by April the G614 substitution reached 80% in Uruguay. In general, the percentage of genomes in South America with this substitution augmented nearly to 100% from March to April (Fig. 2B).

**Figure 2.**
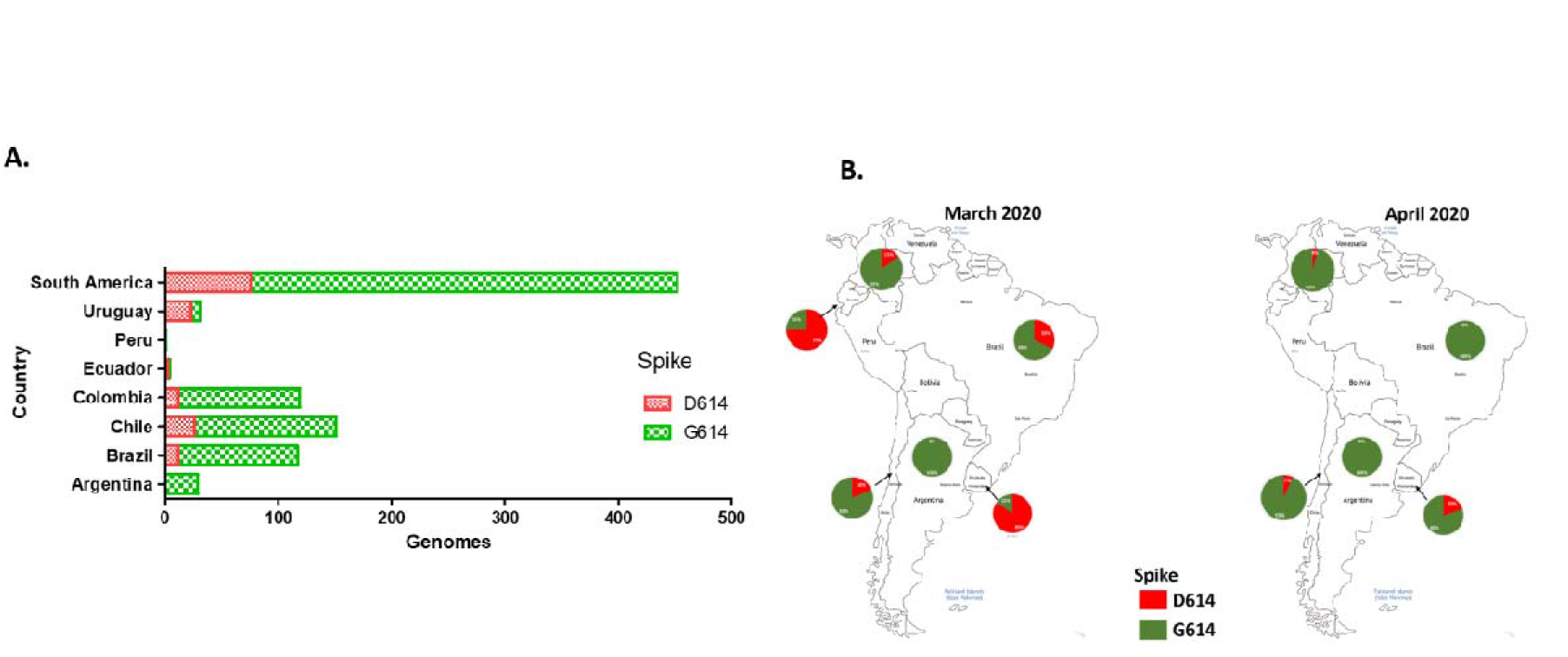
Spatiotemporal distribution of the D614G substitution in the Spike protein of SARS-CoV-2 in South America. **A**. Distribution of D614/G614 substitutions in SARS-CoV-2 genomes reported as of Jun 3th 2020 in South American countries **B**. Spatiotemporal distribution of D614/G614 substitutions in SARS-CoV-2 genomes sequenced between March and April, 2020 in South America, according with the sample collection date.

Non-synonymous substitutions R203K and G204R, which are the hallmarks of the B.1.1 lineage, were the most frequent in the N protein of South American sequences. Both substitutions were frequent in Argentina and Brazil with 55% and 74% of the reported sequences respectively (Fig. 3A). In Ecuador and Chile the frequency of these substitutions was about 20%, while in Uruguay the frequency was similar to Colombia. Furthermore, the proportion of genomes with this double substitution augmented in Chile, Argentina and Brazil from March to April. In contrast, this proportion increased slightly in Colombia and Uruguay, and remained below 20% (Fig. 3B).

**Figure 3.**
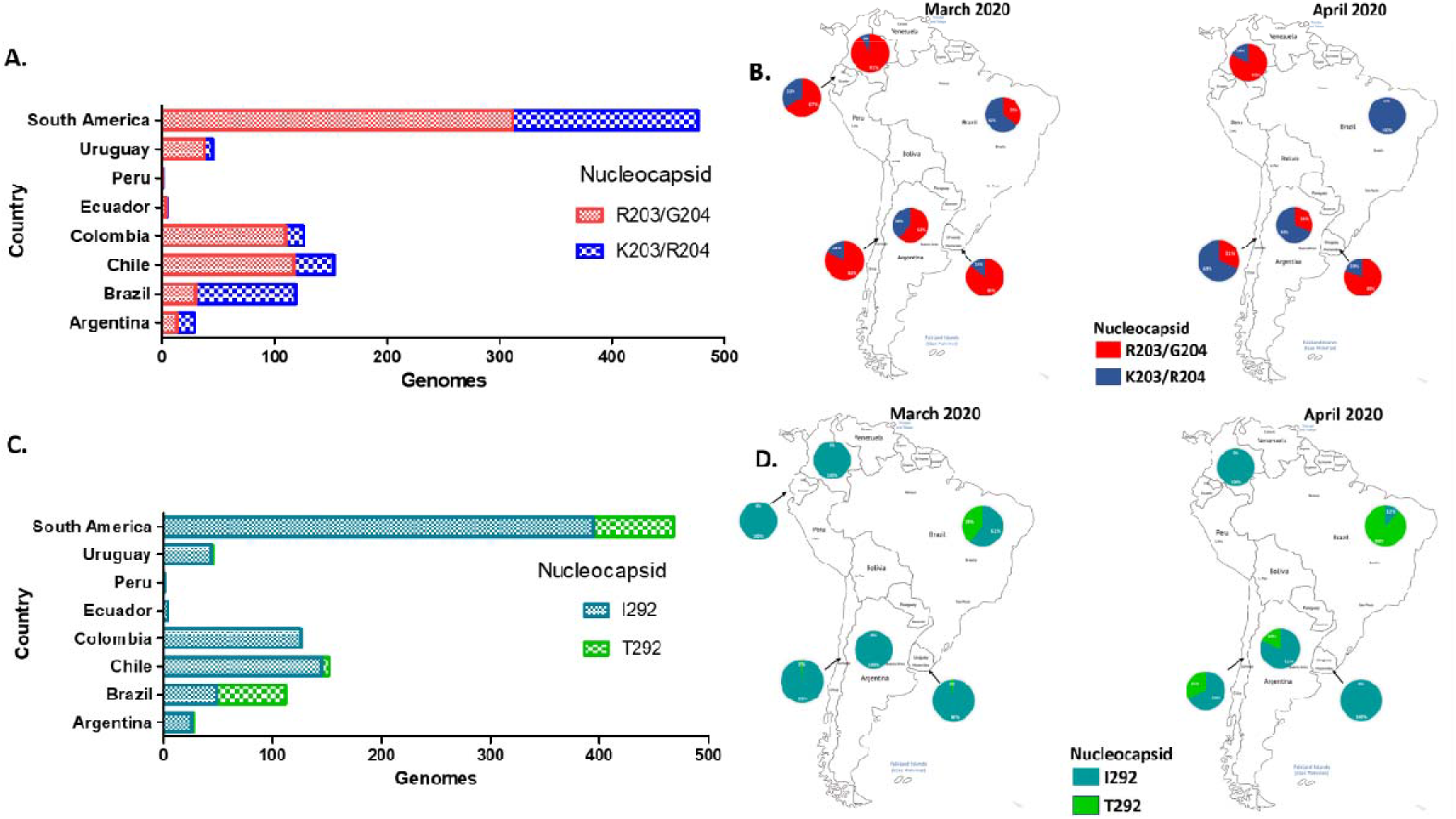
Spatiotemporal distribution of the most frequent substitutions in the Nucleocapsid protein of SARS-CoV-2 in South America. **A**. Distribution of R203/G204 and K203/R204 substitutions in SARS-CoV-2 genomes reported as of Jun 3th, 2020 in South American countries. **B**. Spatiotemporal of R203/G204 and K203/R204 substitutions in SARS-CoV-2 genomes sequenced between March and April, 2020 in South America, according with the sample collection date. **C**. Distribution of I292/T292 substitutions of nucleocapsid protein in SARS-CoV-2 genomes reported as of Jun 3th, 2020 in South American countries. **D**. Distribution of I292 and T292 substitutions in SARS-CoV-2 genomes sequenced between March and April, 2020 in South America, according with the sample collection date.

The substitution I292T in the N protein was rare in Argentina (10.7%), Chile (4.6%) and Uruguay (2.2%); and absent in Colombia, Peru and Ecuador. In contrast, this substitution was very frequent in Brazil (56.3%) (Fig. 3C). The spatiotemporal distribution pattern of this substitution was similar to that of R203K and G204R, increasing from March to April in Chile, Argentina and Brazil in contrast to Colombia and Uruguay where this substitution was almost absent in genomes registered on April (Fig. 3D).

## 4. Discussion

The first COVID-19 case in Colombia was confirmed on March 6, 2020, from a traveler who entered the country from Italy on February 26, 2020 (EPI_ISL_418262). By June 11, 2020, a total 43,810 confirmed cases and 1,505 deaths have been reported (INS, 2020). This study evidenced the presence of the D614G substitution in the S protein in 89.6% (112/125) of Colombian SARS-CoV-2 sequences, by April 27, 2020, while the first introduced cases presented the conserved position reported in the Wuhan-Hu-1 reference genome (Bhattacharyya et al., 2020).

D614G was detected in 85% of sequences being present in most of the South American countries with available genomic information. Several studies have suggested a potential role of the D614G substitution in increase the virus infectivity (Becerra-Flores and Cardozo, 2020; Korber et al., 2020; Nakashima, 2020), transmissibility (Bhattacharyya et al., 2020; Brufsky, 2020), mortality rate and immune system evasion (Kim et al., 2020); however, it was not possible to rule out the association with founder effects.

On the other hand, the co-occurrence of R203K and G204R substitutions in the N protein, was identified in 34% of South American sequences. The B.1.1 lineage is defined by the three-nucleotide mutation in 2 adjacent codons leading to the two consecutive amino acid changes in the N protein (Bartolini et al., 2020; Rambaut et al., 2020), while most of amino acid changes evidenced in the S and N proteins cannot be directly related to a specific lineage (Bhattacharyya et al., 2020; Korber et al., 2020).

This lineage has been reported in samples from travelers with connection to Italy (Gupta and Mandal), also observed in the first confirmed case of SARS-CoV-2 in Colombia (EPI_ISL_418262) and another patient with travel connection to Spain (EPI_ISL_456149) (Table 1). Furthermore, multiple countries outside Italy have reported this lineage among their samples including, Belgium, Switzerland, Vietnam, India, Nigeria and Mexico, demonstrating a wide distribution worldwide (Gupta and Mandal).

RNA viruses are known to possess high substitution rates compared to DNA viruses, leading to high genetic variability and the rapid action of evolutionary mechanisms of natural selection and genetic drift (Li et al., 2020; Tang et al., 2020). Despite some evolutionary changes may be in fact adaptive, it is important to be careful with conclusions in the absence of an experimental model to evaluate the impact of every mutation in the virus phenotype and virus-host interaction (Villabona-Arenas et al., 2020).

The S and N proteins are the most widely used for serological assays, there are 138 FDA-approved serological tests of which only 24% specifically report the screened antigen of these, 39% use the S and 42% use N, and 18% use both (Supplementary Table S3). Recombinant proteins or synthetic peptides of SARS-CoV-2 are widely explored as alternatives to be used in serological tests and therapeutics against SARS-CoV-2 and related Betacoronavirus (Du et al., 2009; Jacofsky et al., 2020), considering that S and N proteins are the major immunogenic proteins of SARS and MERS coronavirus and the first choice for producing recombinant antigens (Yan et al., 2020).

## 5. Conclusion

Amino acid changes were found in the S and N proteins of SARS-CoV-2 circulating in South America, the most frequent being D614G in S, R203K-G204R and I292T in N. It is necessary to continue with genomic surveillance of changes in these proteins during the SARS-CoV-2 pandemic, even more considering that these proteins are the most commonly used in serological and molecular tests. The identification of nucleotide substitutions, amino acid changes and their frequencies in circulating viruses, can be useful for public health decision-making, including vaccine design efforts, design of SARS-CoV-2 diagnostic tests, and therapeutic compounds.

## Data Availability

The authors confirm that the data supporting the findings of this study are available within the article [and/or] its supplementary material.
Raw data were generated at Instituto Nacional de Salud - Unidad de Seguenciacion Analisis Genomico
The data that support the findings of this study are available on request from the corresponding author upon reasonable request.

https://www.gisaid.org/

## 6. Acknowledgements

The authors thank the National Laboratory Network for routine virologic surveillance of SARS-CoV-2 in Colombia. The authors thank all the Colombian and foreign researchers who deposited genomes in GISAID’s EpiFlu (TM) Database contributing to genomic diversity and phylogenetic relationship of SARS-CoV-2.

---

**Supplementary Table S1**. Amino acid substitutions in the Spike protein of SARS-CoV-2 genomes from South America. Location of the substitutions was estimated based on the reference genome NC_045512.2. ^?^ Indeterminate amino acids.

**Supplementary Table S2**. Amino acid substitutions in the Nucleocapsid protein of SARS-CoV-2 genomes from South America. Location of the substitutions was estimated based on the reference genome NC_045512.2. ^?^ Indeterminate amino acids.

**Supplementary Table S3**. Technical details of 138 FDA approved serological Test for anti-SARS-Cov-2 antibody detection.

